# Literacy and mental health: a feasibility and acceptability study among women in a low-income urban setting in North India

**DOI:** 10.64898/2026.09.28.26363834

**Authors:** P.P Shukla, D. Roy, N. Yadav, P.J. Schlüter, K. Mathias

## Abstract

**Introduction:** Literacy is a determinant of mental health, wellbeing, and social participation. Although literacy interventions improve educational and socioeconomic outcomes, their effects on mental health and social participation are poorly evidenced. We aimed to assess the feasibility and acceptability of a group-based literacy and numeracy intervention and explore its effects on literacy, mental health, numeracy, wellbeing, and social participation. We also estimated implementation costs.

**Methods:** We conducted a mixed-methods study using an uncontrolled pre–post design among women in low- income urban communities in Lucknow, India. A total of 115 women were recruited to a 17-week group literacy intervention using the Accelerating Learning for All literacy intervention. Outcomes measured at baseline, midline, and endline used literacy and numeracy assessments and validated scales of mental distress, wellbeing, and social participation. Quantitative data were analysed using multilevel mixed-effects models. Qualitative interviews and focus groups were analysed thematically. Costs were calculated per participant.

**Results:** The intervention was feasible and acceptable with functional recruitment of 71% (82/115), 74/82 participants (90%) completed it (retention), and mean attendance of 69% of scheduled sessions. In multilevel mixed-effects models, mean literacy scores increased by 29.5 (95% CI: 23.6, 35.3) between baseline and endline, while mean mental distress (SRQ20) decreased by 3.4 (95% CI: 2.2, 4.6), mean mental wellbeing (WEMWBS) improved by 2.6 (95% CI: 0.1, 5.1), while social exclusion (P scale) declined by 4.1 (95% CI: 1.6, 6.5). Qualitatively, participants described greater confidence, autonomy, financial literacy, and social support. Implementation enablers included local facilitators.

Implementation cost was US$25 per participant.

**Conclusion:** This low-cost literacy intervention was feasible and acceptable for women in low-income urban communities in India. Findings provide preliminary evidence of improvements in literacy, mental health, and social participation. Controlled studies are needed to establish causal effects, long-term outcomes, and scalability.

**What is already known on this topic:** Literacy is a recognised upstream determinant of mental health through its influence on education, social participation, and health literacy. There is almost no evidence examining whether literacy interventions for women can directly improve mental health and wellbeing, particularly among highly disadvantaged populations. Further, little is known about the feasibility and effectiveness of brief literacy interventions in low-income, disadvantaged urban settings, where women with limited or no literacy skills are disproportionately concentrated.

**What this study adds:** This study shows that a brief literacy intervention among low-income women in informal urban North India is both feasible and acceptable when implemented by local community members. It shows this can be implemented with modest financial cost (around US$25 per person). It shows that participation in the intervention was associated with improved social participation, mental health and mental wellbeing.

**How the study might affect research, practice or policy:** Having demonstrated feasibility and acceptability, together with the promising associations observed in this study, it provides a strong rationale for rigorous randomised controlled trials to establish causality and determine whether this low-cost intervention can deliver meaningful and sustainable improvements in mental health and wellbeing at scale.

## Introduction

Literacy is a key determinant of physical and mental health (1,2) and evidence shows that improved literacy increases equality, child survival and the ability to move out of poverty (3). In an Indian setting, a 2013 study found that literacy was a stronger predictor of health than income inequality or poverty (4).

Low literacy is likely to be an upstream cause of poor health outcomes in India where the adult literacy rate officially stands at 74% (5), while a 2015 study found a functional literacy rate of just 65% (6). Women across India are much less literate than men, reflecting the dominant patriarchy which disadvantages women. In Uttar Pradesh where this study is set, the 2011 census reported literacy rates of 59% among women and 79% among men (5). These disparities are further exacerbated by intersectional structural disadvantage. Earlier, we conducted a study that showed only 10% of rural women aged 51-60 years from scheduled castes in Lucknow district were literate, underscoring the cumulative impact for people experiencing multiple social determinants (6).

Low literacy is associated with poorer health outcomes, particularly poorer mental health. People who cannot read experience higher rates of social isolation, anxiety, poor self-esteem and stigma (7). Low literacy has also been associated with higher rates of depression and other common mental illnesses (8–12) and post-traumatic stress disorder (PTSD) (13). Conversely, when people become literate, this influences other health determinants such as opportunities for employment, income, agency and oral communication skills (14). A recent randomised controlled study of rural women in Uttar Pradesh showed that women who became literate were more actively involved in their children’s education and in promoting family health and hygiene (15).

Evidence shows that adult illiteracy can be addressed through brief interventions, as short as several months (16,17). Yet while studies have suggested that health literacy is protective of mental health and wellbeing (18,19), we have not found other studies examining a literacy intervention for the possible impacts on mental health and wellbeing.

Group literacy classes offer a possible approach to implementation as a cost-effective way to scale delivery of teaching and because they can strengthen other health determinants such as offering new friendships, increased social capital, mutual social support and opportunities for personal development as well as literacy (20). Participation in a psychosocial support group strengthens mental health by supporting participants to increase social networks, foster new peer friendships and increase social support (21–23). Financial literacy is another key determinant of health that has widespread acceptance across low-income communities in South Asia through micro-credit and savings groups known as “self-help groups” (SHGs). These groups typically require participants to hold a level of literacy and numeracy for financial literacy that includes skills in budget planning, savings, autonomy over digital financial transactions and a bank account. Financial literacy is often included as a key skill in mental health interventions, with associated outcomes of greater autonomy, agency and household financial security for low-income households (24,25).

Recognising that literacy can be addressed by brief interventions (16,17), and that groups can additionally promote psychosocial wellbeing, this study aimed to implement and evaluate a group literacy intervention for women with three research aims: firstly, to evaluate the feasibility and acceptability of the intervention and associated measures; secondly, to evaluate preliminary effects of the intervention on the primary outcomes of literacy and mental health, and secondary outcomes of numeracy, wellbeing and social participation; and thirdly, to assess the cost of implementation.

## Methods

### Design

This mixed-methods study used quantitative methods to explore demographics, acceptability and feasibility and pre-post measures at baseline, midline and endline and qualitative methods to explore implementation, feasibility, acceptability and outcomes.

### Setting

Our study was set in a low-income informal urban community, ‘Nai Basti’, comprising one thousand households and around 5000 people in Lucknow, Uttar Pradesh, India. In Nai Basti, people live in makeshift houses on both sides of a railway line and under the Daliganj bridge with limited legal access to electricity and running water. Around 90% of residents are Muslim and the remaining 10% are Hindu and are primarily from the scheduled caste community (the most oppressed caste group).

Implementation was led by Dignity, Education, Vision International Sansthan (DEVI), a non-profit working for foundational literacy and numeracy across India. Women facilitators were selected with these inclusion criteria: had completed at least nine years of schooling, resided in Nai Basti, and were aged 40 years or less. Seven women aged 21–40 years were chosen and six had previous experience as literacy facilitators for children. Facilitators attended a four-hour training on adult learning and group facilitation techniques. Implementers coached and supported facilitators with fortnightly meetings (e.g. discussing how to address challenges in implementation) and connected daily on WhatsApp.

### Participant recruitment

We recruited participants pragmatically, recognising the limited funding we had for implementation. We had seven facilitators and based on previous experience, believed they could teach up to 15 women per class with one facilitator able to teach two classes, thus, we proposed to recruit 15 × 8 women, totalling 120 women. Lanes were randomly selected within Nai Basti, and facilitators knocked on every door to invite women meeting inclusion criteria to participate, accepting a maximum of two women from any single household until they had recruited 15 women. Inclusion criteria were: women aged over 15 years; scoring less than 70% in the literacy test (26); expected to reside in Nai Basti for the duration of the intervention; and willing to join daily classes for 1 to 1.5 hours.

### Procedure

We used the Accelerating Learning for All (ALfA) pedagogy developed by DEVI to teach foundational literacy and numeracy (26). This uses pictures which learners recognise and decode for their first sounds and then blend to form words, approaches that are supported by evidence as effective in teaching adult literacy (17). To teach numeracy, learners use concrete objects (e.g., ice-cream sticks = 10, matchsticks = 1) to represent numbers and solve questions. We also provided reading materials like storybooks, recipes and brochures on topics like cervical screening. Financial literacy was taught weekly using structured modules (27).

Classes ran six days a week from June to October 2025 for 17 weeks with occasional festival holidays. Class times were mutually agreed upon by learners and facilitators, and classes were held within a 200 m radius of participants’ homes, in a shared community space or home. Facilitators were present for 90 minutes daily, but women often came and went (e.g., to attend to children). The routine of classes included homework review, literacy modules, and practice in reading, writing and numeracy in pairs. Storytelling, folk songs, drawing and games were also used. Refreshments were provided weekly.

### Feasibility and acceptability

To evaluate acceptability (perception that the intervention was agreeable and culturally fitting) we measured: a) attendance (a paper register kept by facilitators); b) qualitatively evaluated engagement with the psychometric measures; and, c) programme satisfaction from the perspective of implementers, facilitators and participants.

We evaluated feasibility (the extent to which the intervention was practically able to be implemented in this context) by considering: a) recruitment (number of people who met inclusion criteria who agreed to participate and attended two or more sessions); b) individual and group retention (rates of participation and retention among women and groups through completion of the intervention); and, c) fidelity to intervention (reviewing daily timestamped photographs posted by facilitators on the WhatsApp group as well as using a structured observation format in unannounced weekly visits to each group and attendance registers). Recognising the considerable challenges to engagement in this highly disadvantaged population, we prespecified recruitment, attendance, and retention rates of at least 65% as benchmarks of feasibility and acceptability.

### Data collection

Nursing students and women graduates collected quantitative data. DR, NY and PPS conducted a half-day training for data collectors in ethics, consent processes and the psychometric scales followed by a rehearsal of procedures in the field. Due to participants’ limited literacy, a trained data collector verbally administered all quantitative measures in Hindi and recorded participants’ responses. Women participants met in their groups and serially completed data forms with data collectors in June (baseline), August (midline) and October (endline) 2025. We omitted the midline data collection of the P scale to reduce participant burden based on feedback from facilitators following baseline data collection.

### Quantitative outcome measures

We used National Mental Health Survey data fields for socio-demographic data which included fields of caste, gender, religion and age as well as type of housing, presence of a toilet, filtered water and an electricity meter (28). We measured literacy by combining three standardised and validated tools developed in and used widely across India to assess reading, writing and numeracy/financial literacy summing to a total of 100 marks. The reading test (50 marks) assessed participants’ reading using four items using the Annual Status of Education report (ASER) test to categorise each participant’s reading proficiency into one of five levels ranging from letter recognition to reading a paragraph(29–31). The writing (30 marks) and numeracy tools (20 marks) are components of the Foundational Literacy and Numeracy Assessment Test. These tools were designed for adults with low literacy and were developed by the Ministry of Education under the ULLAS / Nav Bharat Saaksharta Karyakram(32) adult literacy programme. Across reading, writing and numeracy, the range of scores was 0 to 100. Women scoring more than 70 at registration were deemed literate and excluded from the intervention.

We used these three psychometric scales: mental distress was measured using the SRQ20 scale; social inclusion, using the Participation scale (P scale), and wellbeing assessed using the Warwick-Edinburgh Mental Wellbeing Scale (WEMWBS). The SRQ20 scale was developed in low-resource settings (33), validated in South Asia (34,35), and consists of 20 items which sum from 0-20. Higher SRQ20 scores indicate greater mental distress. The P scale (36,37) was developed and validated in South Asia, consists of 18 items, and yields scores from 0-28. Higher scores indicate greater social exclusion (37). The WEMWBS is a widely employed 14-item instrument, has been validated among women in informal urban settings in India (24), and yields a total score of 14-70. Higher WEMWBS scores denote greater mental wellbeing (38). To avoid issues with missingness, the scales were checked immediately after completion, and data collectors sat with participants to fill gaps in the few cases where missing items were recorded. Quantitative tools were each reviewed by the research associate with data collectors from the target community to check for comprehension. In a few instances tools were slightly modified to reflect the Urdu-flavoured Hindi idiom in the study setting by the research associate. The modified version was back-translated by an independent Urdu and Hindi speaker and pilot-tested to assure comprehension.

### Qualitative

Qualitative data were collected by PPS with KM supporting at several time points throughout the intervention. We interviewed implementers, facilitators, women learners and data collectors (9 individual participants). Two focus group discussions (FGDs) were conducted with facilitators and three with learners (a total of 35 participants).

The semi-structured guides used for interviews and FGDs sought to explore experiences of facilitators and learners, including enablers and challenges of participation and questions like ‘what made it easy/difficult to participate in classes?’ and ‘what changed for you after joining literacy classes?’ Copies of interview guides are available in Supplementary materials. We also explored the strategies facilitators employed to address implementation challenges and possible mechanisms and outcomes of participation. Recordings were translated and transcribed in English by PPS. Transcripts were read and inductively coded by KM and PPS using Open Code 4.03(39) using the six-stage reflexive thematic analysis of (a) familiarization, (b) initial code generation, (c) theme construction, (d) revision, (e) theme definition, and (f) report preparation. Themes emerged inductively from data and were grouped into the headings deductively derived from the evaluation framework (40).

### Statistical analysis

Reporting of the quantitative findings was informed by the STROBE guidelines (41) and included comparison of the demographics of participants who completed vs. non-completers using Fisher’s exact test. Following summary statistics of measures, we analysed multilevel mixed-effects models for each primary outcome variable, with participants nested within their facilitator groups which were treated as random effects. Facilitator and participant intraclass correlations (ICCs) were reported, with mean changes between assessment periods and 95% confidence intervals (CIs). Residual checks were undertaken for all models, and only important violations were noted. Statistical analyses were conducted using Stata SE version 18.0 (StataCorp, College Station, Texas, USA), and a two-sided α=0.05 was used to determine significance.

### Cost

We recorded all costs linked to implementation and tallied these at the end of the grant implementation. These were divided by total participants who commenced the intervention (n=82) to calculate a cost per participant in INR.

### Public participation

This study was co-designed with DEVI Sansthan team members residing in Nai Basti. Women from target communities were employed as facilitators and contributed to the study design and implementation by adapting survey instruments and psychometric scales to local language and context, advising on recruitment, and shaping delivery strategies to maximise participation, including locating groups within 200 m of participants’ homes. Although they were not involved in selecting psychometric measures, community members contributed to the design of qualitative components, including the processes and guides for focus groups. A plain-language Hindi summary of the study findings was shared with facilitators and participants.

### Ethics

Ethics approval was provided by both the University of Canterbury, New Zealand (HREC 2025/16) and KGMU, India (144^th^ ECM IIA/P14). The clinical trials registry of India identification was CTRI/2025/06/089429.

### Positionality

PPS, DR and NY are Indian nationals while KM is an Overseas Citizen of India and PJS is a New Zealand citizen. All researchers except PJS worked in Hindi. While none of the authors were ongoing residents in the study site, several of them have spent many weeks working and at times living in similar settings which helped give insight into the living conditions and implementation challenges.

## Results

### Participants

Of 115 women screened, 103 met inclusion criteria and consented to baseline data collection (90% of those invited). However, 21 of these women withdrew from the programme almost immediately, attending 0 or 1 literacy sessions. Anecdotal reports suggested that some had expected financial or other rewards that were not provided. This left 82 participants from the original 115 women invited, a functional recruitment of 71%. After starting the intervention, 8 participants withdrew in the first six weeks due to family challenges (e.g., caring for children or husband limiting participation) resulting in 74 women completing the intervention (90% retention). Participation in the study is presented in a PRISMA diagram in Figure 1.d

**Figure 1.**
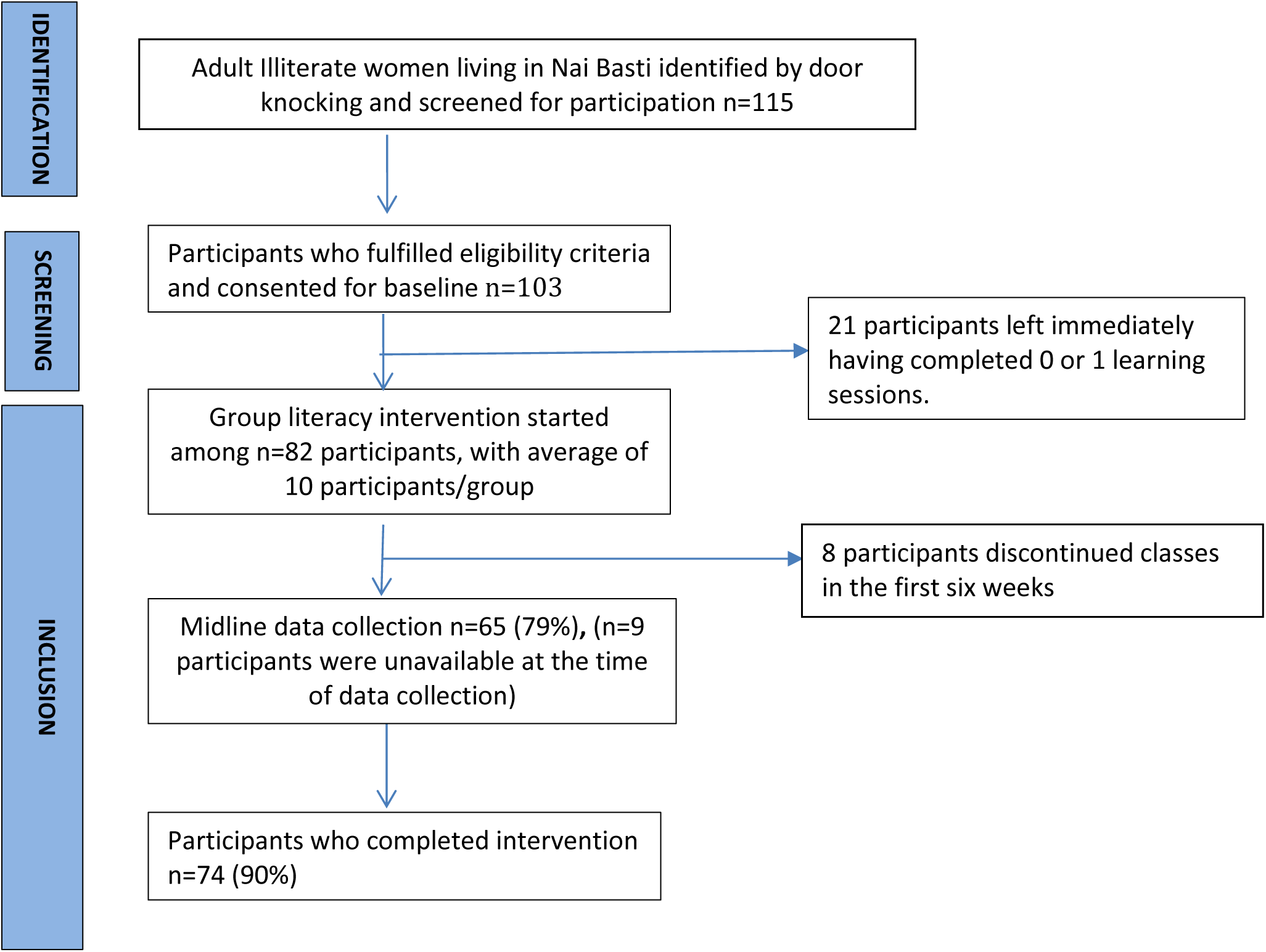
PRISMA diagram showing participants in the intervention and the three data collection points.

### Participant characteristics

Sociodemographic characteristics are presented in Table 1. Most participants were 18 to 59 years, married, Muslim, from disadvantaged communities, with little formal schooling and often ran out of money.

**Table 1.** Sociodemographic characteristics of recruited women at the baseline survey.

|  | Participated |  | Withdrew after<br>0 or 1 session |  | p-value* |
| --- | --- | --- | --- | --- | --- |
|  | n | (%) | n | (%) |  |
| **Age (years)** |  |  |  |  | 0.84 |
| 15-17 | 4 | (5) | 2 | (10) |  |
| 18-59 | 73 | (89) | 18 | (86) |  |
| ≥60 | 5 | (6) | 1 | (5) |  |
| **Marital status** |  |  |  |  | 0.49 |
| Married | 69 | (84) | 16 | (76) |  |
| Divorced | 0 | (0) | 0 | (0) |  |
| Widow | 4 | (5) | 2 | (10) |  |
| Unmarried | 9 | (11) | 3 | (14) |  |
| **Religion** |  |  |  |  | 0.68 |
| Muslim | 74 | (90) | 20 | (95) |  |
| Hindu | 8 | (10) | 1 | (5) |  |
| Other | 0 | (0) | 0 | (0) |  |
| **Caste category** |  |  |  |  | 0.67 |
| General | 12 | (15) | 5 | (24) |  |
| OBC | 59 | (72) | 13 | (62) |  |
| SC/ST | 5 | (6) | 1 | (5) |  |
| Not known | 6 | (7) | 2 | (10) |  |
| **Type of house and facilities** |  |  |  |  | 0.08 |
| Kuccha house (made of temporary materials like tarpaulin) | 47 | (57) | 17 | (81) |  |
| Semi-Kuccha house (concrete floor with low-quality roof and walls) | 11 | (13) | 0 | (0) |  |
| Pucca (made with permanent materials) | 24 | (29) | 4 | (19) |  |
| Presence of toilet | 50 | (61) | 10 | (48) | 0.46 |
| Presence of filtered water | 10 | (12) | 12 | (57) | <0.001 |
| Presence of electricity with meter | 62 | (76) | 11 | (52) | 0.11 |

\*Calculated using Fisher's exact test.

Comparing participants (n=82) against those who left almost immediately after recruitment (n=21), there was no significant difference in sociodemographic characteristics except for having a water filter within the home (12% vs. 57%, p<0.001).

### Attendance and retention

Table 2 presents mean attendance and retention for groups over the maximum possible of 102 literacy sessions. Seven of eight groups continued to operate throughout the intervention. Group 8 which was small (n=4 women), dissolved due to challenges in the outdoor location by the railway tracks and one woman joined group 2 (n=1) while three joined group 4 (n=3). The attendance records of these participants were merged with their new groups for simplicity. Attendance at classes held indoors ranged from 71%–78% and was notably poorer for classes set in open (outdoor) settings. Facilitator in group 1 managed higher attendance in an open space, perhaps due to her strong community linkages. Initially eight classes were formed. One class held adjacent to a railway track (separate from group 3) was discontinued after two months due to low participant attendance and four women dropped out. Key barriers to attendance identified by women and facilitators were linked to physical circumstances including lack of shade, the open areas becoming very muddy in monsoon, constrained spaces and interference by men in the area. Women across all groups attended a mean of 69% of classes. Across classes, retention of women ranged from 56% to 100% with a mean retention rate of 90%.

**Table 2.**
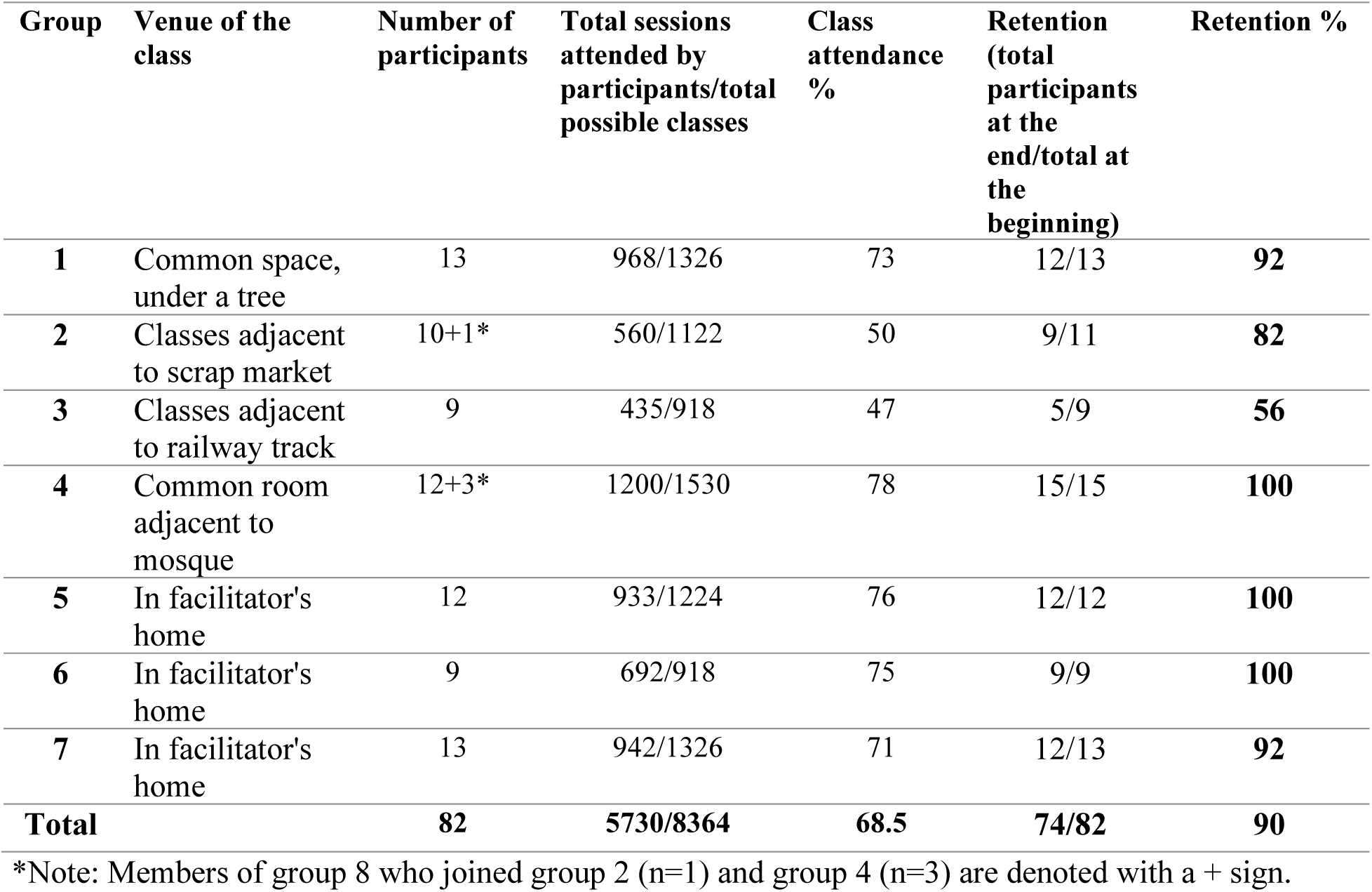
Summary of attendance and retention by group for those who participated (n=82).

Fidelity was monitored through qualitative observations by the intervention coach, focusing on group facilitation and application of ALfA pedagogical principles. Observations suggested that facilitators largely adhered to the intervention model, effectively using instructional materials and peer coaching to support learning.

### Primary variables

Summary statistics of the primary outcome variables at baseline, midline and endline are presented in Table 3. The mean literacy score increased from baseline to midline (p<0.001) and again from midline to endline (p=0.01) representing continued improved literacy. The mean WEMWBS (mental wellbeing) increased from baseline to midline (p=0.03) but did not change significantly between midline and endline (p=0.84). Conversely, SRQ20 (mental distress) did not significantly decrease from baseline to midline (p=0.08) but did so from midline to endline (p<0.001). P scale scores decreased from baseline to endline (p=0.001).

**Table 3.** Summary statistics and multilevel mixed-effects model estimates of the primary outcome variables over time: Literacy; SRQ20 score; WEMWBS; and P scale score.

| Multilevel mixed-effects model estimates |  |  |  |
| --- | --- | --- | --- |
|  | N | mean | (95% CI) |
| Literacy score |  |  |  |
| Baseline | 82 | 25.5 | (19.2, 31.9) |
| Midline | 63 | 46.6 | (39.8, 53.5) |
| Endline | 74 | 55.0 | (48.4, 61.5) |
| SRQ20 mental distress score |  |  |  |
| Baseline | 82 | 12.5 | (11.2, 14.0) |
| Midline | 63 | 11.5 | (10.0, 13.0) |
| Endline | 74 | 9.2 | (7.8, 10.6) |
| WEMWBS wellbeing score |  |  |  |
| Baseline | 81 | 50.7 | (48.9, 52.6) |
| Midline | 62 | 53.6 | (51.5, 55.7) |
| Endline | 74 | 53.3 | (51.4, 55.3) |
| P scale score |  |  |  |
| Baseline | 82 | 12.0 | (10.2, 13.9) |
| Endline | 74 | 8.0 | (6.0, 9.9) |

### Primary outcomes

Multilevel mixed-effects models with participants nested within facilitator groups were used to assess changes in primary outcomes over time. Model-estimated means and 95% CIs are shown in Table 3 and Figure 2. Literacy scores increased significantly from baseline to midline (mean change: 21.1 points, 95% CI: 14.9, 27.3; p=0.001) and from midline to endline (mean change: 8.3 points, 95% CI: 2.0, 14.7; p=0.01). Mental distress non-significantly decreased from baseline to midline (mean change: −1.1, 95% CI: −2.4, 0.1; p=0.08) but decreased significantly from midline to endline (mean change: −2.3, 95% CI: −3.5, −1.0; p<0.001), while mental wellbeing improved from baseline to midline (mean change: 2.9, 95% CI: 0.3, 5.5; p=0.03) but showed no further change thereafter (mean change: −0.3, 95% CI: −2.9, 2.4; p=0.84). Social exclusion also declined significantly between baseline and endline (mean change: −4.1, 95% CI: −6.5, −1.6; p<0.001). Facilitator-level clustering was low (ICC: <0.01-0.10), whereas participant-level clustering ranged from low to moderate (ICC: 0.02-0.56). No important model assumption violations were detected.

**Figure 2.**
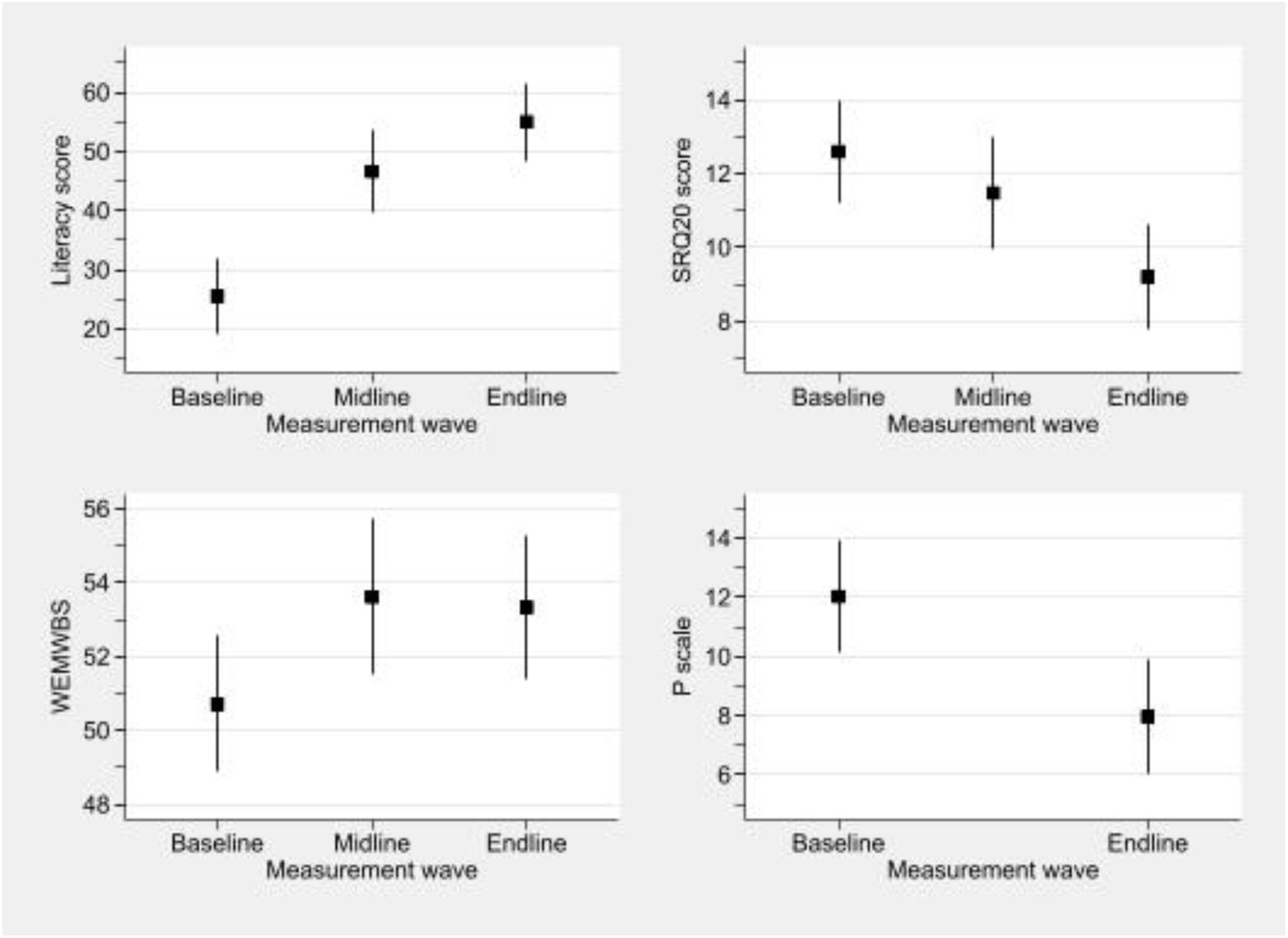
Multilevel mixed-effects model mean estimates and associated 95% CIs for literacy score, WEMWBS, SRQ20 score, and P scale score.

### Feasibility

#### Qualitative findings

The qualitative findings are presented under the headings of acceptability, feasibility of research components and feasibility of implementation followed by implementation adaptations and outcomes.

#### Acceptability of intervention

Participants described that they felt comfortable and at home with the group intervention format, although there were some initial challenges with the small number of women from a privileged caste expressing that they were unwilling to sit with others from less advantaged castes.

Strict, gendered norms such as restrictions on mobility, burden of household work and caregiving were often barriers to women attending the classes. A woman who stopped attending after a few weeks reflected,

> *“When men of the house return (home), I have to serve them food. I also have kids to look after. It is impossible to attend classes, even though I want to.”*

Another challenge was conflicts among two or more participants of a group meaning they would stop attending the classes temporarily. These conflicts generally resolved after group facilitators counselled the participants. Women described facilitators as ‘like us’ and many were known as local residents, using the same dialect, dress and cultural norms as women participants.

Group facilitators generally reported a high level of satisfaction with the intervention and felt well trained and supported. However, facilitators of groups held in open settings described frustrations with interruptions, low attendance and difficult physical situations (heat from direct sun, rain and mud) and women in these groups also described the physical conditions as challenging.

Implementers described the intervention as feasible and acknowledged that this was facilitated by previous experience as literacy teachers for children. They proposed that greater training, coaching and support would be required if implementing this intervention with facilitators new to literacy teaching.

#### Feasibility of research components

Women and data collectors found the psychometric measures easy to comprehend and responded readily. Women were able to focus and respond well to the psychometric test when it was filled in a location where they were undisturbed by their family members, children or peers. Baseline data collection took approximately 60 minutes per participant which led to some respondents feeling fatigued. Subsequent phases required only 30 minutes of time per participant as sociodemographic details were not required which was more feasible. Researchers reviewed the quality of survey data collected by nursing students and women graduates. Both groups collected data that were of high quality with a high level of survey completion. Importantly, as locals to the area, they used approaches and language that was culturally sensitive and appropriate.

### Feasibility of implementation - challenges

#### Patriarchy and social hierarchies limited freedom of movement and participation

Patriarchal gender norms that positioned women’s roles as within the household and diminished the perceived value of female literacy were frequently reported by facilitators and learners. Many women required their husbands’ permission to attend classes, some participated covertly, and others were restricted from travelling independently to and from sessions. For example, one woman indicated that decision-making lay with her husband, saying:

> *“My husband is educated and he never stops me from learning. But he wants me home, when he comes in the afternoon to have lunch and take a nap.”*

#### Socioeconomic deprivation limited participation

Women in the most deprived housing and communities experienced more difficult learning environments which were linked to the open-air settings’ physical conditions (like rain and mud). Those with young children found attending classes difficult due to interruptions. The poorest women also found participation challenging (for example, those living next to the railway tracks were reluctant to leave their children unattended) and, in the most disadvantaged communities, unemployed men disturbed classes, as one facilitator described:

> *“I am no longer taking classes along the railway tracks. Men over there interfere. They look inside and ask me innumerable questions, disturbing the class and we feel uncomfortable”.*

### Feasibility of implementation - enablers

#### Creative activities and refreshments increased participation

Facilitators reported higher attendance on days when snacks were provided. Additional strategies to enhance participation included escorting women from their homes to overcome mobility restrictions and incorporating engaging activities such as games, drawing, cooking, storytelling, and local excursions.

#### Peer friendship strengthened learning

Women reported that the group supported social connectedness, providing opportunities to meet neighbours and build relationships. Learning was often collaborative, with participants supporting each other’s reading, while peer-led competition encouraged engagement and progress. Women also supported one another in walking to classes.

#### Facilitators with shared lived experiences strengthened trust and participation

Facilitators shared socio-demographic identities with women learners which increased a sense of trust and engagement. Learners described that facilitators spoke and lived just like them which meant they understood their circumstances, and adapted classes as required. Previous experience in teaching literacy to children is likely to have assisted facilitators in teaching literacy to adults.

#### Agile problem-solving addressed disruptions

Although learning conditions were highly challenging in disadvantaged settings, facilitators adapted to complex social and environmental circumstances. Strategies included splitting classes across locations when interpersonal tensions arose, relocating sessions to shaded outdoor spaces or participants’ homes when needed, and providing iced water during hot weather. Observations highlighted the difficulties of open-air classes, with classes sometimes held on small mats amid mud, rubbish, and frequent interruptions from children, men, and animals.

Implementers highlighted the value of locating classes close to participants’ homes, ongoing facilitator coaching to support problem-solving and maintain fidelity to the ALfA methodology, and facilitators’ capacity to adapt delivery to learners’ circumstances. WhatsApp also served as a valuable tool for accountability, peer support, collective problem-solving, and sharing both challenges and successes.

### Outcomes

#### Increased financial literacy

Learners and facilitators reported increased confidence in managing household finances, including shopping calculations, budgeting, and saving. Facilitators also observed healthy competition among learners, motivating some women to reduce their discretionary spending and save more consistently. One woman in her mid 30’s described how this helped her with shopping:

> “Before, when I used to buy rations from a shop, I couldn’t calculate the total, but now I can. I even pose questions to the shopkeeper.”

#### Improved confidence and autonomy

Facilitators described that women developed greater confidence and self-esteem in classes. Another participant described this:

> *“Yes, many things have changed. Earlier, we used to hesitate to talk to people outside, we felt shy or scared. But now our confidence has grown. Now we don’t feel afraid to talk or ask questions. Earlier, we hesitated to speak in the presence of educated people, but not anymore.”*

Women also reported greater autonomy, such as in attending hospital appointments, travelling on public transport and completing forms independently. One woman described that she was able to describe health issues to her doctor such as the date of her last menstrual period, overcoming previous embarrassment about not knowing such details.

#### Increased peer friendships and social support

Learners and facilitators described that the group supported formation of new social relationships in both indoor and outdoor settings. Although women knew each other as neighbours, the group provided an opportunity for deeper friendships as outlined below:

> *“We discuss and share what the facilitator has taught us. We also share our sorrows, issues, conflicts at home with each other. I have known everyone since childhood here. Earlier we were not meeting our friends daily. Now that is possible. The best part is, despite being married, we meet and greet each other.”*

#### Improved wellbeing to join a bigger world

Learners described the classes as opportunities to engage in new activities, including drawing, cooking, and embroidery. These experiences broadened their horizons, strengthened their sense of connection beyond the home, and contributed to improved wellbeing. One learner described this as follows:

> *“Classes are also an opportunity for us to learn many things that we were not aware of. The class is like a window to the world”.*

### Fidelity of the intervention

Facilitators posted the timestamped photographs of classes for 100% of sessions. Based on facilitator notes in attendance registers, mean duration of literacy classes was 90 minutes. Observations during unscheduled visits by the intervention supervisor were supplemented by discussions with facilitators and indicated that literacy classes and activities were conducted consistently.

### Cost of intervention

The total implementation cost comprising training and coaching of facilitators, facilitator remuneration, learning materials and refreshments was INR 195,000 or US$2,050. Divided by 82 (the number of women who commenced classes) equated to approximately INR 2,400 or US$25 per participant.

## Discussion

This feasibility study provides evidence that a 17-week group-based literacy intervention for women living in a low-income urban community in northern India was acceptable and feasible when classes were held indoors. The classes held outside had low attendance and one of four outside classes was discontinued suggesting these were not feasible settings for the intervention. Women attended a mean of 70 of 102 classes (69%), despite many living in highly disadvantaged settings such as homes under bridges and under tarpaulin.

Group facilitation was enabled by local women who shared participants’ language, religion, and community context and acted as cultural mediators, enhancing trust, attendance, engagement, cultural safety, and connection (42,43). Other enablers included setting learning sessions at times that suited women, and adjusting timings and locations during festivals or school holidays, features which have strengthened implementation in other low-income settings (44).

Although this was primarily an intervention study and not powered for a large quantitative study, our findings suggest that participation in this intervention was associated with significant improvements in literacy and wellbeing and reductions in social exclusion and mental distress. The low facilitator-level ICCs suggest good consistency in programme delivery across facilitators, with little evidence that facilitator effects substantially influenced participant outcomes. By comparison, participant-level ICCs ranged from low to moderate, indicating that some outcomes exhibited considerable within-person dependence and that stable individual differences accounted for an appreciable proportion of the observed variation. Together, these findings imply that variability in outcomes was driven primarily by participant-level factors rather than differences between facilitators.

These group-level dynamics are likely to have supported engagement and may partly explain the high acceptability and retention observed. At the same time, the presence of clustering highlights the importance of accounting for group-level effects in both analysis and the design of future, controlled studies.

This study contributes novel evidence to the limited literature examining a brief literacy intervention as mental health–promoting. Other studies have described the association of low literacy with poor mental health, but we have not found other studies evaluating literacy interventions in terms of mental health and social connection outcomes (2,45). While we did not formally explore the mechanisms by which literacy influences mental health, our qualitative outcomes reported greater agency, self-efficacy and wellbeing, with women describing classes opening a window to a bigger world. This aligns with wider literature showing that literacy is correlated with improved self-confidence, agency, health literacy, involvement in social activities (46), communication and ability to access information technology including social media (47). Literacy also influences other health determinants such as social capital and employment (48). Theories proposed to support the ways that literacy protects people from poor mental health include that it supports greater health knowledge, fertility choices, self-efficacy and lifestyle choices (such as influencing choices in cigarette smoking) (1). Apart from these explicit mechanisms, education can improve health indirectly through opportunity for higher income, increased learning ability, skill sets and accumulated social capital (18).

Our findings point to several opportunities to strengthen implementation. Recruitment should clearly communicate both the benefits and expectations of participation, including the absence of financial incentives. Delivering classes in indoor venues may reduce distractions and improve learning conditions, while childcare provision could enable attendance. Developing more relevant, adult-oriented reading materials may further increase participant engagement and retention.

Key research implications are to evaluate whether this intervention is effective using a randomised controlled methodology; secondly, to formally explore the mechanisms that might be operating for literacy to increase mental health and/or protect from mental ill-health and to consider whether this intervention is feasible and effective in other populations and settings (such as women in rural areas).

Our findings point to the possibility of adult literacy programmes as a cost-effective intervention that impacts multiple determinants of wellbeing, self-efficacy and mental health. While adult literacy does not sit clearly in either education or health sectors, this feasibility study suggests it should be explored as an intersectoral social investment which can potentially improve population mental health for women and their families to reduce inequities, increase agency and strengthen social participation.

### Methodological considerations

Our study has several strengths in its design. A strength of our approach was the statistical modelling, which allowed us to estimate changes in quantitative measures within the observational design.

Quantitative controlled studies typically require rigid interventions and research design which would have been difficult in this first iteration and a disadvantaged context (45). By using qualitative methods, we could collect and analyse nuanced data linked to context, which allowed us to document micro-adaptations and intervention changes as the study progressed. Secondly, by interviewing multiple stakeholders, including women, facilitators, and implementers, we could triangulate findings to provide verification of data. We also provided detailed descriptions of the context and implementation which strengthens the potential for scaling and generalisability (external validity).

There are also limitations to the methodology. Without a control group, we could not infer causality in the outcomes noted. Secondly, 21 women who participated in baseline data collection chose to exit the intervention at inception, due to a misunderstanding where they believed there would be financial benefits for participation. However, we noted no statistical difference in the demographic characteristics of this group to the intervention group except the difference in the presence of a water filter in the home, which potentially is a spurious type I error.

## Conclusion

This study shows that this low-budget group literacy intervention implemented by community members in local meeting spaces in Nai Basti, Lucknow, was both feasible and acceptable. Further, our findings suggest that the intervention is associated with improved scores in literacy, mental health and wellbeing, and social participation. This study identifies barriers and enablers for implementing this literacy intervention in a low-income urban setting as well as pragmatic, iteratively developed solutions to implementation challenges. A large-scale randomised controlled trial in diverse settings will be needed to determine the generalisability and effectiveness of this group literacy intervention to promote mental health and social inclusion.

## Authorship

PPS led research implementation, including data collection, and, with KM, completed the qualitative analysis. DR led ethics and trial registration and, with NY, supported data collection and analysis. PJS and KM applied for research funding and co-designed the study, and PJS conducted the quantitative data analysis. PPS and KM wrote the first draft. All authors contributed to subsequent drafts of the paper and approved the final version.

## Data Availability

All data produced in the present study are available upon reasonable request to the author

## Acknowledgements

The authors express appreciation to Dr Sunita Gandhi, founder of DEVI Sansthan and developer of ALfA. We also appreciate the support of her entire team. In addition, we thank Afreen, Anjum, Heena, Ishrat, Mariyam, Nisha and Zeba, the community literacy facilitators and data collection team, which included nursing students and data collectors from the community, for their energy and time, and the generous participation of all the women learners in interviews, data collection and focus group meetings. DEVI Sansthan supported the implementation by providing materials and training for the volunteers.

## Notes

### Competing Interest Statement

The authors have declared no competing interest.

### Clinical Trial

CTRI/2025/06/089429

### Author Declarations

Human Research Ethics Committee of University of Canterbury, New Zealand gave ethical approval for this work. The Institutional Ethics Committee of Kings George Medical University of Lucknow , India gave ethical approval for this work

## References

1. Zhang Q. The cost of illiteracy: A causal inference study on how illiteracy affects physical and mental health. Health Educ J. 2021 Feb;80(1):54–66. doi:10.1177/0017896920949894

2. Hunn L, Teague B, Fisher P. Literacy and mental health across the globe: a systematic review. Ment Health Soc Incl. 2023 Dec 1;27(4):392–406. doi:10.1108/MHSI-09-2022-0064

3. Azevedo J, Rogers H, Akmal M, Cloutier M, Ding E. The State of Global Learning Poverty. Washington (DC): World Bank; 2022.

4. Rajan K, Kennedy J, King L. Is wealthier always healthier in poor countries? The health implications of income, inequality, poverty, and literacy in India. Soc Sci Med. 2013 Jul;88:98–107. doi:10.1016/j.socscimed.2013.04.004

5. Office of the Registrar General, Government of India. Census 2011: Lucknow District[Internet]. Office of the Registrar General, Government of India. Available: http://www.census2011.co.in/census/district/528-lucknow.html

6. Gandhi S, Mathias K, Seefeldt L, Delaney. The Inconvenient Truth of Illiteracy Inequality. Econ Polit Wkly. 2021;56(34):61.

7. Easton P, Entwistle VA, Williams B. How the stigma of low literacy can impair patient-professional spoken interactions and affect health: insights from a qualitative investigation. BMC Health Serv Res. 2013 Dec;13(1):319. doi:10.1186/1472-6963-13-319

8. Kohli C. Prevalence of Unrecognised Depression Among Outpatient Department Attendees of A Rural Hospital in Delhi, India. J Clin Diagn Res. 2013. doi:10.7860/JCDR/2013/6449.3358

9. Mathias K, Goicolea I, Kermode M, Singh L, Shidhaye R, Sebastian MS. Cross-sectional study of depression and help-seeking in Uttarakhand, North India. BMJ Open. 2015 Nov;5(11):e008992. doi:10.1136/bmjopen-2015-008992

10. Nguyen T, Tran T, Tran H, Tran T, Fisher J. The burden of clinically significant symptoms of common and severe mental disorders among adults in Vietnam: a population-based cross-sectional survey. BMC Public Health. 2019 Dec;19(1):1173. doi:10.1186/s12889-019-7513-7

11. Gupta C, Arora M, Gupta R, Akhtar N, Langer B, Kumari R, et al. Prevalence and correlates of depression in a rural adult population in Northwest India. J Fam Med Prim Care. 2020;9(1):151. doi:10.4103/jfmpc.jfmpc_656_19

12. Firdaus G. Mental well-being of migrants in urban center of India: Analyzing the role of social environment. Indian J Psychiatry. 2017;59(2):164. doi:10.4103/psychiatry.IndianJPsychiatry_272_15

13. Adhikari Baral I, K.C B. Post traumatic stress disorder and coping strategies among adult survivors of earthquake, Nepal. BMC Psychiatry. 2019 Dec;19(1):118. doi:10.1186/s12888-019-2090-y

14. Akello JA, Lutwama-Rukundo E, Musiimenta P. Functional Adult Literacy: An Alternative Gateway to Grassroots Women’s Improved Income Generation in Lango Subregion, Northern Uganda. Adult Educ Q. 2017 May;67(2):79–96. doi:10.1177/0741713616685143

15. Deshpande A, Ksoll C, Maertens A, Varghese V. The Impact of an Adult Education Program for Mothers: Evidence from India. Econ Dev Cult Change. 2023 Jan 1;71(2):709–51. doi:10.1086/715116

16. Kolinsky R, Carvalho C, Leite I, Franco A, Morais J. How to Teach Fully Illiterate Adults to Read. In: Perin D, editor. The Wiley Handbook of Adult Literacy [Internet]. 1st ed. Wiley; 2019 [cited 2026 Jul 7]. p. 407–28. Available from: https://onlinelibrary.wiley.com/doi/10.1002/9781119261407.ch19 doi:10.1002/9781119261407.ch19

17. Abadzi H. Adult literacy: A review of implementation experience [Internet]. Washington DC, USA: World Bank; 2003. Available from: https://mavmatrix.uta.edu/context/curriculuminstruction_facpubs/article/1050/type/native/viewcontent

18. Berkman ND, Sheridan SL, Donahue KE, Halpern DJ, Crotty K. Low Health Literacy and Health Outcomes: An Updated Systematic Review. Ann Intern Med. 2011 Jul 19;155(2):97–107. doi:10.7326/0003-4819-155-2-201107190-00005

19. Chevalier A, Feinstein L. Sheepskin or Prozac: The Causal Effect of Education on Mental Health. Inst Study Labor IZABonn [Internet]. 2006;(Discussion Paper No. 2231). Available from: https://www.econstor.eu/bitstream/10419/33928/1/515785318.pdf

20. Prins E, Toso BW, Schafft KA. “It Feels Like a Little Family to Me”: Social Interaction and Support Among Women in Adult Education and Family Literacy. Adult Educ Q. 2009 Aug;59(4):335–52. doi:10.1177/0741713609331705

21. Barbui C, Purgato M, Abdulmalik J, Acarturk C, Eaton J, Gastaldon C, et al. Efficacy of psychosocial interventions for mental health outcomes in low-income and middle-income countries: an umbrella review. Lancet Psychiatry. 2020 Feb;7(2):162–72. doi:10.1016/S2215-0366(19)30511-5

22. Gailits N, Mathias K, Nouvet E, Pillai P, Schwartz L. Women’s freedom of movement and participation in psychosocial support groups: qualitative study in northern India. BMC Public Health. 2019 Dec;19(1):725. doi:10.1186/s12889-019-7019-3

23. Mathias K, Pillai P, Gailits N. How Do Psychosocial Support Groups in North India Support Collective Action for Mental Health? A Qualitative Study Using a Caring Methodology. Asia Pac Viewp. 2025 Aug;66(2):165–76. doi:10.1111/apv.12451

24. Kanougiya S, Daruwalla N, Osrin D. Mental health on two continua: mental wellbeing and common mental disorders in a community-based cross-sectional study with women in urban informal settlements in India. BMC Womens Health. 2024 Oct 9;24(1):555. doi:10.1186/s12905-024-03389-1

25. Pal RK. Financial stress: is it self-driven or fate-driven? J Soc Welf Manag. 2025;17(1). doi:10.21088/jswm.2454.7964.11125.5

26. Gandhi S. ALfA fast and holistic booster Hindi Reading & Writing [Internet]. Lucknow: Devi Sansthan.Available: https://dignityeducation.org/ [Accessed 1 May, 2025]

27. The Diksha Program for Financial Literacy & Debt Management [Internet]. Parinaam foundation; 2014. Available: https://unituslabs.org/wp-content/uploads/mf-innovations/Ujjivan-Parinaam-Annexure-I-Diksha-Financial-Literacy-Program-Trainer-Manual.pdf

28. Gururaj G, Varghese M, Benegal V, Rao G, Pathak K, Singh L. National Mental Health Survey of India, 2015–16: Summary. Bengaluru: NIMHANS, Ministry of Health and Family Welfare, Government of India; 2016.

29. Vagh SB. Validating the ASER testing tools: Comparisons with reading fluency measures and the Read India measures. [Unpublished report]. 2012. [Accessed 18 August, 2026]

30. Kumar D, Sunder N, Sabates Aysa R, Wadhwa W. Improving children’s foundational learning through community-school participation: Experimental evidence from rural India. Labour Econ. 2024 Dec;91:102615. doi:10.1016/j.labeco.2024.102615

31. Richards J. Extending ASER as an assessment tool throughout South Asia. Int J Educ Dev. 2024 Nov;111:103152. doi:10.1016/j.ijedudev.2024.103152

32. National Council of Educational Research and Training. Cell for national centre for literacy NCERT [Internet]. NCERT; 2026. Available: https://ncert.nic.in/cncl/test_papers.php [Accessed 18 August, 2026]

33. Beusenberg M, Orley JH. A user’s guide to the Self Reporting Questionnaire (SRQ). Geneva: World Health Organization Division of Mental Health; 1994.

34. Husain N, Gater R, Tomenson B, Creed F. Comparison of the Personal Health Questionnaire and the Self Reporting Questionnaire in rural Pakistan. JPMA J Pak Med Assoc. 2006 Aug;56(8):366–70. PubMed PMID: 16967789.

35. Patel V, Araya R, Chowdhary N, King M, Kirkwood B, Nayak S, et al. Detecting common mental disorders in primary care in India: a comparison of five screening questionnaires. Psychol Med. 2008 Feb;38(2):221–8. doi:10.1017/S0033291707002334

36. Stevelink SAM, Hoekstra T, Nardi SMT, Van Der Zee CH, Banstola N, Premkumar R, et al. Development and structural validation of a shortened version of the Participation Scale. Disabil Rehabil. 2012 Sep;34(19):1596–607. doi:10.3109/09638288.2012.656793

37. Van Brakel WH, Anderson AM, Mutatkar RK, Bakirtzief Z, Nicholls PG, Raju MS, et al. The Participation Scale: Measuring a key concept in public health. Disabil Rehabil. 2006 Jan;28(4):193–203. doi:10.1080/09638280500192785

38. Grover S, Dua D. Hindi Translation and Validation of Scales for Subjective Well-being, Locus of Control and Spiritual Well-being. Indian J Psychol Med. 2021 Nov;43(6):508–15. doi:10.1177/0253717620956443

39. Open Code 4.03 [Internet]. Sweden: Department of Public Health and Clinical Medicine, Umea University; 2018. Available: http://www.phmed.umu.se/english/divisions/epidemiology/research/open-code/?languageId=1 [Accessed 4 July, 2025]

40. Craig P, Dieppe P, Macintyre S, Michie S, Nazareth I, Petticrew M. Developing and evaluating complex interventions: the new Medical Research Council guidance. BMJ. 2008 Sep 29;a1655. doi:10.1136/bmj.a1655

41. Von Elm E, Altman DG, Egger M, Pocock SJ, Gøtzsche PC, Vandenbroucke JP. The Strengthening the Reporting of Observational Studies in Epidemiology (STROBE) statement: guidelines for reporting observational studies. J Clin Epidemiol. 2008 Apr;61(4):344–9. doi:10.1016/j.jclinepi.2007.11.008

42. Mackintosh NJ, Davis RE, Easter A, Rayment-Jones H, Sevdalis N, Wilson S, et al. Interventions to increase patient and family involvement in escalation of care for acute life-threatening illness in community health and hospital settings. Cochrane Consumers and Communication Group, editor. Cochrane Database Syst Rev. 2020 Dec 8;2020(12). doi:10.1002/14651858.CD012829.pub2

43. Kearns N, Randle J, Taggart A, Tizzoni S, Quinn A, Watene J. Reimagining health in Porirua: a community-led approach to hauora. N Z Med J. 2025 Oct 24;138(1624):13–26. doi:10.26635/6965.7078

44. Mishra A, Singh AK, Parida SP, Pradhan SK, Nair J. Understanding Community Participation in Rural Health Care: A Participatory Learning and Action Approach. Front Public Health. 2022 Jun 6;10:860792. doi:10.3389/fpubh.2022.860792

45. Haeri-Mehrizi A, Mohammadi S, Rafifar S, Sadighi J, Kermani RM, Rostami R, et al. Health literacy and mental health: a national cross-sectional inquiry. Sci Rep. 2024 Jun 13;14(1):13639. doi:10.1038/s41598-024-64656-7

46. Olomukoro CO, Adelore OO. Political Empowerment of Women through Literacy Education Programmes in EDO and Delta States, Nigeria. J Adult Contin Educ. 2015 Nov;21(2):3–23. doi:10.7227/JACE.21.2.2

47. Allatt G. Adult literacy education and social inclusion. Centre for Research in Education Inclusion & Diversity, University of Edinburgh; 2017. Available: https://eprints.hud.ac.uk/id/eprint/32505/ [Accessed 7 July, 2025]

48. Stromquist, N. P. (2009). Literacy and empowerment: A contribution to the debate. Background study commissioned in the framework of the United Nations Literacy Decade, UNESCO, Paris; 2009.

